# An uncorrected hematology analyzer change distorts blood-count measures of aging and inflammation in NHANES: A cross-sectional study

**DOI:** 10.64898/2026.11.26.26369040

**Authors:** Tiago V. Barreira, Kylie K. Harmon, Eduardo C. Costa, Matthew K. Armstrong, Kevin S. Heffernan

**Author notes:** Corresponding author (TVB). Department of Human Performance, Falk College of Sport, Syracuse University, 150 Irving Ave, Syracuse, NY 13244.

## Abstract

**Background:** Composite measures built from routine blood counts, including phenotypic age, the red cell distribution width (RDW)–to–albumin ratio (RAR) and inflammation ratios, appear in more than a thousand National Health and Nutrition Examination Survey (NHANES) studies, most pooling many cycles. NHANES replaced its hematology analyzer in 2013–2014 and noted shifts in several analytes, but stored whole blood cannot be re-assayed, so no bridging equations exist. This study quantified the resulting distortion and derived correction factors where possible.

**Methods and findings:** In 11 NHANES cycles (1999–2023; *n* = 88,943), changes in 12 blood-count measures were estimated at all 10 adjacent-cycle boundaries in adolescents, in whom true two-year change is not expected; phenotypic age and ratio measures were examined in 48,961 adults. At the analyzer boundary, RDW rose 0.72 percentage points (95% CI 0.58–0.86; 5.7%; *p* < 0.001) and white blood cell count (WBC) 0.42 × 10³/µL (0.05–0.79; 6.3%; *p* = 0.026), for both the largest change at any boundary. In adults, the transition added 2.63 years (2.13–3.13; *p* < 0.001) of apparent phenotypic age acceleration, creating an apparent post-2012 surge in biological aging and nearly doubling the share of later participants in the top pooled quartile (35% vs 19%); subtracting the adolescent RDW and WBC changes from 2013–2014 onward removed 92.0% of this increase (residual 0.21 years; *p* = 0.40), whereas chemistry placebo corrections changed it by ≤7.1%. Among ratio measures, RAR rose 6.5% (*p* < 0.001) and the platelet-to-lymphocyte ratio fell 4.8% (*p* = 0.020), neither remaining significant after correction (*p* ≥ 0.12); the neutrophil-to-lymphocyte ratio, in which WBC cancels, did not change (*p* = 0.97). The main limitation is reliance on an adolescent referent rather than split samples.

**Conclusions:** An uncorrected analyzer change adds 2.6 years of artifactual phenotypic aging from 2013–2014 onward. Subtracting 0.72 percentage points from RDW and 0.42 × 10³/µL from WBC corrects these two inputs; red cell indices and platelets should not be pooled across cycles. Analysts should check each cycle’s laboratory documentation for every variable before pooling NHANES data.

**Author summary:** *Why was this study done?:*

- More than a thousand studies use blood-count measures from the US National Health and Nutrition Examination Survey (NHANES), such as phenotypic age and inflammation ratios, and most pool many survey cycles.
- NHANES replaced its hematology analyzer in 2013–2014 and noted shifts in several blood counts, but because whole blood cannot be stored for split-sample testing, no bridging equations exist and studies pool the data without adjustment.

*What did the researchers do and find?:*

- Changes in 12 blood-count measures were estimated at all 10 boundaries between survey cycles from 1999 to 2023 in adolescents, in whom true two-year change is not expected, and their consequences were tested in 48,961 adults.
- Red cell distribution width and white blood cell count rose about 6% at the analyzer change and at no other boundary, constant across percentiles; in adults, this added 2.6 years of apparent phenotypic age, produced an apparent post-2012 surge in biological aging, and nearly doubled the share of adults examined from 2013 onward who fell in the top phenotypic age quartile when cycles were pooled (35% vs 19%).
- Subtracting the two adolescent shifts from post-2013 values removed 92% of the increase in phenotypic age and the shifts in the ratio measures that contain them, whereas the same procedure applied to chemistry analytes had little effect.

*What do these findings mean?:*

- Multicycle NHANES studies spanning 2013 that compare levels, trends or pooled quantiles of measures containing red cell distribution width or white blood cell count are distorted and may need reexamination.
- Two fixed values correct these two inputs; ratios in which white blood cell count cancels need no adjustment; red cell indices and platelets should not be pooled across cycles.
- The correction rests on an adolescent referent rather than split samples, but the step it removes was the same across young, middle-aged and older adults.

## Introduction

The National Health and Nutrition Examination Survey (NHANES) makes over two decades of laboratory data publicly available, and open-source code and artificial intelligence tools now facilitate multicycle studies without the need to open each cycle’s laboratory documentation [1]. Composite measures calculated from routine blood tests, including phenotypic age [2,3] and ratio measures such as the red cell distribution width (RDW)–to–albumin ratio (RAR), the platelet-to-lymphocyte ratio (PLR), the systemic immune-inflammation index (SII) and the neutrophil-to-lymphocyte ratio (NLR), are widely used in studies of aging, inflammation and mortality, and hematological data are increasingly used to build new aging clocks [4,5]. A PubMed search of titles and abstracts through September 2026 returned 1,164 NHANES studies using these measures, 90% of them published since 2024 [6,7].

NHANES replaced its hematology analyzer (Beckman Coulter MAXM with the DxH 800) between the 2011–2012 and 2013–2014 cycles. When a biochemistry platform changes, the National Center for Health Statistics (NCHS) runs split-sample comparisons and publishes bridging equations [8]; for the complete blood count, however, method-comparison studies on stored specimens were not possible [9]. NCHS therefore documented that RDW, white blood cell count (WBC), hematocrit, mean corpuscular hemoglobin (MCH) and its concentration (MCHC) shifted at that point, “indicating possible method issues,” and advised researchers to “consider these possible method changes when analyzing trends” [9], but could offer no correction. None of the pooled-cycle analyses reviewed for this study accounted for the change, and the analysis toolkits used to compute these measures take the released values as comparable across cycles [10,11].

Without an empirical estimate of the shift, analysts cannot tell a correctable instrument step from the ordinary drift of a measure across cycles, nor judge how much it distorts the composite measures built on it. The purpose of this study was to quantify the effect of the 2013–2014 analyzer change on each blood-count measure and on the composite measures built from them, to evaluate its consequences for pooled analyses, and to derive correction factors where possible.

## Methods

### Study design and data

This cross-sectional analysis of repeated NHANES cycles is reported following the Strengthening the Reporting of Observational Studies in Epidemiology (STROBE) guideline for cross-sectional studies [12] (S1 STROBE Checklist). No protocol or analysis plan was registered. The primary comparison, the change in each blood-count measure between the 2011–2012 and 2013–2014 cycles in adolescents, was defined at the outset, after the NCHS documentation of the analyzer change was identified; the series across all adjacent-cycle boundaries, the percentile test of correction form, the chemistry placebo tests and the quartile analysis were added during the analysis, between August and October 2026, to test the specificity and consequences of that change, and the phlebotomy-weight sensitivity analysis was added in November 2026. Eleven cycles were used, 1999–2000 through August 2021–August 2023, excluding the 2017–March 2020 prepandemic file, which re-releases the 2017–2018 participants. Demographic files (DEMO to DEMO_L), complete blood count files (LAB25, L25_B, L25_C and CBC_D to CBC_L) and standard biochemistry files (LAB18, L40_B, L40_C and BIOPRO_D to BIOPRO_L) were downloaded with the nhanesA package.

### Participants

The analytic frame comprised all examined, nonpregnant participants with valid RDW and WBC (*n* = 88,943), without age restriction. The adolescent referent comprised participants aged 12–19 years (*n* = 2,380 in the 2011–2012 and 2013–2014 cycles combined; 1,983–4,301 per boundary). The adult phenotypic age sample (*n* = 48,961; 9,272 in the 2011–2012 and 2013–2014 cycles) comprised participants aged 20 years or older and below their cycle’s top-coded age (85 years in 1999–2006, 80 years from 2007–2008), after values of the eight inputs more than five standard deviations from their within-cycle mean were set to missing, mirroring the cleaning applied in the BioAge package.

### Change between adjacent cycles

For each measure and each of the 10 adjacent-cycle boundaries, the change is the coefficient on an indicator for the later cycle in a survey-weighted linear regression restricted to the two cycles and to the adolescent referent. Percentage changes divide by the earlier cycle’s weighted mean. A trend model with a step term was not used, because it attributes any departure from the assumed trend to the step. The 2011–2012 to 2013–2014 boundary is termed the analyzer boundary. NCHS indicates that the instruments were replaced during 2012, so part of the 2011–2012 cycle was measured on the new analyzer and the change estimated between the two cycle means is conservative [9]. The adolescent referent follows the reasoning NCHS itself used when attributing a shift in hemoglobin A1c to a method change because it appeared in younger participants, in whom increases would not be expected over a short period.

### Correction rule and form

A measure was considered correctable when NCHS had identified it as affected by the analyzer change [9], its largest change across the 10 boundaries occurred at the analyzer boundary, and the change was similar in size across the distribution. To decide between an additive and a proportional correction, the change was estimated at the 10th, 25th, 50th, 75th and 90th percentiles from survey-weighted quantiles in each cycle, treating the cycles as independent samples; a constant absolute change indicates an additive correction and a constant percentage change a proportional one (S2 Table). NHANES reports RDW and WBC to one decimal place, so the step sizes at each percentile are discrete.

### Phenotypic age

Phenotypic age was computed with the phenoage_calc function of the BioAge package (version 0.1.0) [10,11], trained in NHANES III adults aged 20–84 years and projected onto NHANES 1999–2023 with the fixed fit. Because C-reactive protein was not collected in 2011–2014, the model used albumin, creatinine, glucose, lymphocyte percentage, mean corpuscular volume (MCV), RDW, alkaline phosphatase and WBC with chronological age. Chemistry analytes were adjusted across the 2017–2018 and 2021–2023 laboratory transitions with the NCHS bridging equations, applied as a two-stage chain [8]. Acceleration is the residual of phenotypic age on chronological age in the pooled adult sample. The change in phenotypic age at the analyzer boundary was estimated in adults with chronological age as a covariate, overall and within three age groups (20–39, 40–59 and 60 years or older), with an age-group-by-cycle interaction term to test whether the change differed by age.

### Ratio measures

Four ratio measures were computed from the same frame: RAR (RDW divided by albumin), PLR (platelet count divided by the absolute lymphocyte count), SII (platelet count multiplied by the absolute neutrophil count and divided by the absolute lymphocyte count) and NLR (absolute neutrophil count divided by absolute lymphocyte count). Because each absolute count is WBC multiplied by the corresponding differential percentage, WBC remains in the denominator of PLR but cancels in SII and NLR, and RAR contains RDW directly. The change in each ratio at the analyzer boundary was estimated in adults as a percentage of the 2011–2012 mean.

### Correction applied and placebo tests

For participants examined from 2013–2014 onward, the adolescent change was subtracted from RDW and WBC, all other values were held fixed, and phenotypic age was re-projected through the same fit; the share of the step removed is one minus the ratio of the corrected to the measured change. The same procedure was applied to RDW alone, to WBC alone and, as placebo tests, to each chemistry analyte, for which no platform changed at this boundary. The corrected RDW and WBC values were also used to recompute RAR and PLR in adults. For display, the corrected acceleration series is expressed against the observed age fit, so that it coincides with the measured series before 2013–2014.

### Quartile reassignment

Survey-weighted quartile cut points were computed once in the 11-cycle pooled adult sample and separately within each cycle, and the percentage of adults whose quartile differs between the two assignments is reported.

### Literature count

PubMed was searched on 19 October 2026 for records published through 30 September 2026, with all terms restricted to the title or abstract: (“phenotypic age” OR “PhenoAge” OR “red cell distribution width to albumin ratio” OR “red blood cell distribution width to albumin ratio” OR “RDW to albumin ratio” OR “RDW/albumin” OR “RDW-to-albumin” OR “red cell distribution width-to-albumin” OR “red blood cell distribution width-to-albumin” OR “systemic immune-inflammation index” OR “systemic immune inflammation index” OR “neutrophil-to-lymphocyte ratio” OR “neutrophil to lymphocyte ratio” OR “systemic inflammation response index” OR “platelet-to-lymphocyte ratio” OR “platelet to lymphocyte ratio”) AND (NHANES OR “National Health and Nutrition Examination Survey”). Counts include all article types and were not screened.

### Statistical analysis

All estimates used the two-year mobile examination center weight (WTMEC2YR) for every cycle, including 1999–2002, divided by the number of cycles pooled, with variance from the masked strata and primary sampling units (survey package version 4.5). Repeating the analyses with the phlebotomy weight (WTPH2YR) that NCHS recommends for blood analytes in 2021–2023 left the results essentially unchanged; for example, mean phenotypic age acceleration in 2021–2023 was 1.05 rather than 1.02 years. Each coefficient was tested with a t statistic on design-based degrees of freedom (number of primary sampling units minus number of strata). Tests are two-sided at α = 0.05. No data were excluded beyond the stated eligibility criteria and the five-standard-deviation cleaning rule, and no imputation was performed. Analyses were run in R version 4.5.1 in RStudio 2026.07.1.

### Ethics statement

NHANES protocols were approved by the NCHS Research Ethics Review Board (Protocol #98-12 for 1999–2004; #2005-06 for 2005–2010; #2011-17 for 2011–2012 through October 2017; #2018-01 from October 2017; and #2021-05 for 2021–2023; https://www.cdc.gov/nchs/nhanes/about/erb.html). Adult participants gave written informed consent; for participants younger than 18 years, a parent or guardian gave written permission and the participant gave assent. This analysis of deidentified, publicly available data required no further review.

### Use of artificial intelligence

Two artificial intelligence assistants were used. Claude (Anthropic; Claude Fable 5.1, accessed via claude.ai, September–October 2026) helped write and debug the R analysis code and edit the text, tables and figure. Gemini Pro (Google; Argon, October 2026) was used to review the writing and check reported numbers against the analysis output. All code, outputs and text were reviewed and verified by the authors, who take full responsibility for the content.

## Results

### Blood-count measures

Seven of 12 blood-count measures changed at the analyzer boundary (Fig 1A and Table 1). RDW rose by 0.72 percentage points (95% CI 0.58–0.86; 5.7%; *p* < 0.001) and WBC by 0.42 × 10³/µL (95% CI 0.05–0.79; 6.3%; *p* = 0.026). For both, this was the largest change across the 10 boundaries, and the change was similar across the distribution (10th to 90th percentiles: 0.6–0.8 percentage points for RDW and 0.4–0.5 × 10³/µL for WBC), supporting an additive correction (S2 Table). The RDW change matched the full-sample change reported by NCHS (12.84% to 13.55%) [9]. RDW showed only one other significant change, a 1.3% rise across the pandemic gap between 2017–2018 and 2021–2023, one-quarter the size of the analyzer step (S1 Table).

**Fig 1.**
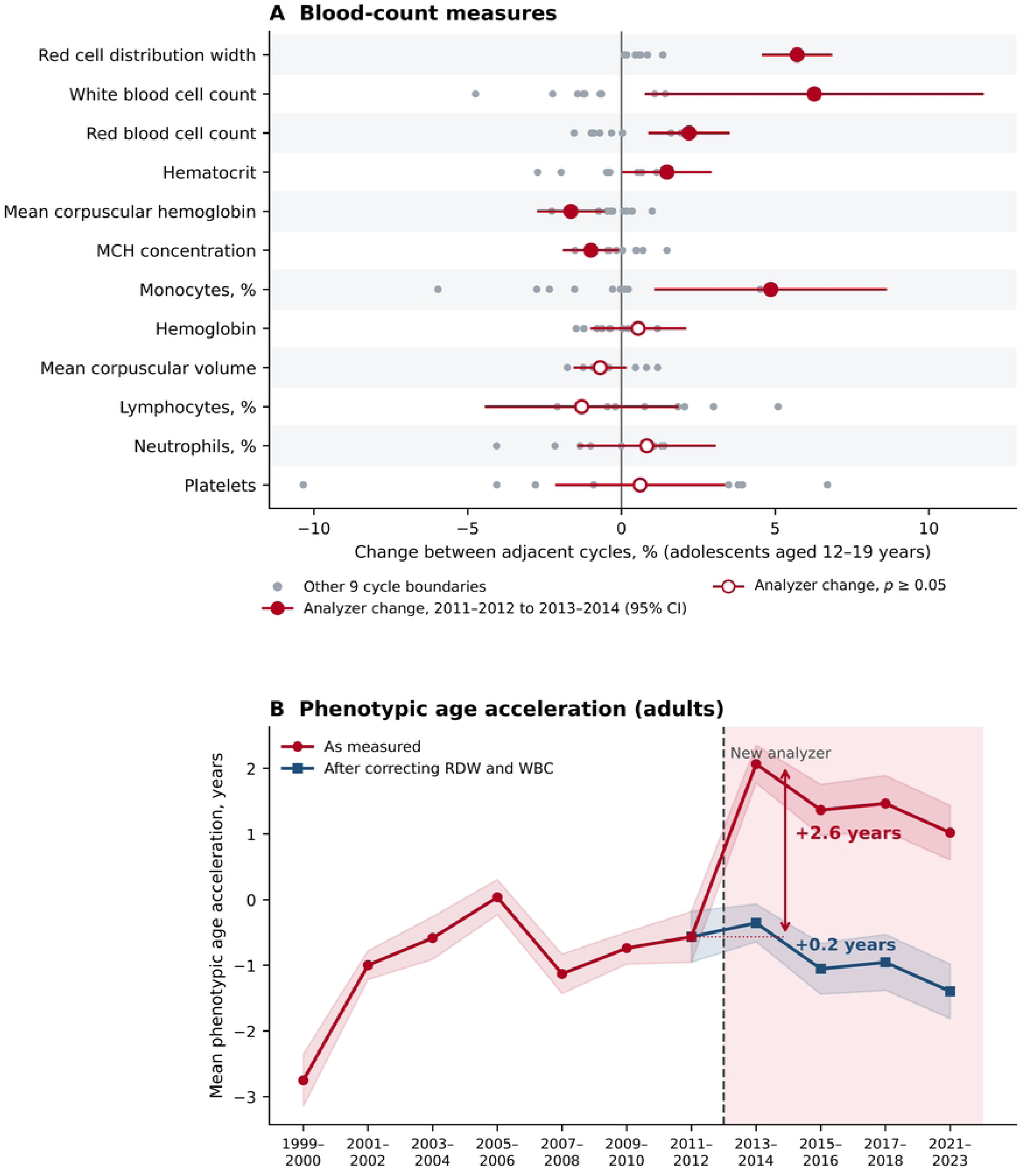
Effect of the 2013–2014 hematology analyzer change on blood-count measures and phenotypic age in NHANES, 1999–2023. (A) Change in the survey-weighted mean of each measure between adjacent cycles among adolescents aged 12–19 years (*n* = 1,983–4,301 per boundary), as a percentage of the earlier cycle. Gray dots, the nine other cycle boundaries; red circles, the analyzer boundary (2011–2012 to 2013–2014) with 95% CI from survey-weighted linear regression; open circles, *p* ≥ 0.05. (B) Mean phenotypic age acceleration by cycle in adults aged 20 years or older (*n* = 48,961), as measured and after subtracting the adolescent changes in RDW and WBC from values measured from 2013–2014 onward; both series are expressed on the same reference and coincide before 2013–2014. Shading, 95% CI; dashed line, the analyzer change; the low 1999–2000 value reflects an earlier chemistry analyzer that is not corrected here. RDW, red cell distribution width; WBC, white blood cell count.

**Table 1.** Change in blood-count measures at the analyzer boundary in adolescents and the correction, if any.

| **Measure** | **Change at the analyzer boundary, % (95% CI)** | **Largest other change, %** | **Correction for 2013–2014 onward\*** |
|---|---|---|---|
| **Largest change at the analyzer boundary: correctable** |  |  |  |
| RDW, % | 5.7 (4.6 to 6.8) | 1.3 | Subtract 0.72 (95% CI 0.58–0.86) |
| WBC, × 10<sup>3</sup>/µL | 6.3 (0.8 to 11.7) | −4.7† | Subtract 0.42 (95% CI 0.05–0.79) |
| **Changed at the analyzer boundary and at other boundaries: not correctable with a single factor** |  |  |  |
| Red blood cell count | 2.2 (0.9 to 3.5) | 1.9 | None |
| Hematocrit‡ | 1.5 (0.1 to 2.9) | −2.7 | None |
| MCH‡ | −1.6 (−2.7 to −0.6) | −2.3 | None |
| MCHC‡ | −1.0 (−1.9 to −0.1) | −1.5 | None |
| Monocytes, % | 4.9 (1.1 to 8.6) | −6.0 | None |
| **No change at the analyzer boundary** |  |  |  |
| Hemoglobin | 0.5 (−1.0 to 2.1) | −1.5 | Not needed |
| MCV | −0.7 (−1.5 to 0.1) | −1.8 | Not needed |
| Lymphocytes, % | −1.3 (−4.4 to 1.8) | 5.1 | Not needed |
| Neutrophils, % | 0.8 (−1.4 to 3.0) | −4.1 | Not needed |
| Platelets | 0.6 (−2.1 to 3.3) | −10.3§ | Not needed§ |

Change is the difference in survey-weighted means between the 2011–2012 and 2013–2014 cycles among adolescents aged 12–19 years (*n* = 2,380), as a percentage of the 2011–2012 mean, with 95% CI from survey-weighted linear regression. ‘Largest other change’ is the largest change at any of the nine other adjacent-cycle boundaries (S1 Table). NCHS reports the following full-sample changes between the same cycles: RDW 12.84% to 13.55%; WBC 7.03 to 7.41 × 10<sup>3</sup>/µL; hematocrit 40.57% to 41.22%; MCH 30.56 to 29.99 pg; MCHC 34.34 to 33.90 g/dL [9].
\* Subtract from values measured from 2013–2014 onward, before computing any composite measure; the change was similar across the distribution (10th, 50th and 90th percentiles: 0.8, 0.6 and 0.8 percentage points for RDW; 0.4, 0.4 and 0.5 × 10<sup>3</sup>/µL for WBC; S2 Table).
† NCHS reports a decline in WBC between 2005–2006 and 2011–2012 [13], which the correction does not address.
‡ Calculated by the analyzer from red blood cell count, MCV and hemoglobin; NCHS attributes their 2013–2014 changes mainly to a single DxH 800 instrument [9].
§ Platelets changed at seven of the nine other boundaries.
MCH, mean corpuscular hemoglobin; MCHC, mean corpuscular hemoglobin concentration; MCV, mean corpuscular volume; RDW, red cell distribution width; WBC, white blood cell count.

Red blood cell count rose by 2.2%, and hematocrit (1.5%), MCH (−1.6%) and MCHC (−1.0%) changed as well. The analyzer calculates these three indices from red blood cell count, MCV and hemoglobin, and with red blood cell count up 2.2% and hemoglobin unchanged (0.5%; *p* = 0.47), their observed changes match those predicted arithmetically (1.5%, −1.7% and −1.0%). Monocyte percentage also changed (4.9%). Each of these five measures, however, changed by a similar or larger amount at another boundary, and NCHS attributes the red cell index changes mainly to a single instrument, whereas the RDW and WBC increases occurred on all three [9]. Hemoglobin, MCV, lymphocyte and neutrophil percentages and platelets did not change at the analyzer boundary, although platelets changed at seven of the nine other boundaries (S1 Table).

### Phenotypic age

In adults, mean phenotypic age acceleration rose by 2.63 years at the analyzer boundary (95% CI 2.13–3.13; *p* < 0.001), similarly across age groups (2.52–2.81 years; interaction *p* = 0.47) (Fig 1B). As measured, acceleration went from −0.57 years in 2011–2012 to 2.07 years in 2013–2014 and remained above 1.0 year through 2021–2023, an apparent post-2012 surge that no earlier boundary approaches. With quartiles cut in the pooled sample rather than within cycle, 26.5% of adults changed phenotypic age quartile. Subtracting the adolescent changes from RDW and WBC for cycles from 2013–2014 onward removed 92.0% of the step, leaving 0.21 years (95% CI −0.29 to 0.71; *p* = 0.40); RDW alone removed 79.8% and WBC alone 12.2%, and the apparent surge disappeared (Fig 1B). As measured, the top pooled quartile held 34.6% of adults examined from 2013 onward and 18.9% of those examined before; after correction, it held 25.3% and 24.8%, and the share of adults changing quartile fell from 26.5% to 11.5%. The same procedure applied to chemistry analytes, for which no platform changed, altered the step by 7.1% or less.

### Ratio measures

The ratio measures followed the analytes they contain (Table 2). RAR, which contains RDW, rose by 6.5% (95% CI 5.4–7.7; *p* < 0.001), and PLR, whose denominator contains WBC, changed by −4.8% (95% CI −8.8 to −0.8; *p* = 0.020), whereas SII (−0.7%; *p* = 0.79) and NLR (−0.1%; *p* = 0.97), in which WBC cancels, did not change. With quartiles cut in the pooled sample rather than within cycle, 30.8% of adults changed RAR quartile, compared with 4.1% for NLR. After correction, the RAR change was 0.9% (*p* = 0.12), the PLR change 1.7% (*p* = 0.39), and the share of adults changing RAR quartile 11.9%.

**Table 2.** Effect of the analyzer change on phenotypic age and the ratio measures in adults, as measured and after correction.

| **Measure** | **Shifted analytes contained** | **Change, as measured** | **Change after correction** | **Adults changing quartile, %** | **Correction** |
|---|---|---|---|---|---|
| Phenotypic age acceleration | RDW, WBC | 2.63 y (*p* < 0.001) | 0.21 y (*p* = 0.40) | 26.5 | Correct RDW and WBC |
| RDW-to-albumin ratio | RDW | +6.5% (*p* < 0.001) | +0.9% (*p* = 0.12) | 30.8 | Correct RDW |
| Platelet-to-lymphocyte ratio | WBC, in denominator | −4.8% (*p* = 0.020) | +1.7% (*p* = 0.39) | 19.2 | Correct WBC* |
| Systemic immune-inflammation index | None; WBC cancels | −0.7% (*p* = 0.79) | — | 13.7 | Not needed* |
| Neutrophil-to-lymphocyte ratio | None; WBC cancels | −0.1% (*p* = 0.97) | — | 4.1 | Not needed |

Adults aged 20 years or older in the 2011–2012 and 2013–2014 cycles (*n* = 9,272); difference between the two cycles, phenotypic age adjusted for chronological age and ratios as a percentage of the 2011–2012 mean; p values from survey-weighted regression. After correction: RDW and WBC measured from 2013–2014 onward reduced by the Table 1 factors before each measure was computed. Adults changing quartile: percentage assigned to a different quartile when quartiles are cut in the 11-cycle pooled sample rather than within cycle. After correction, 11.5% of adults changed phenotypic age quartile and 11.9% changed RAR quartile.
\* Platelet-based measures are not comparable across cycles for reasons unrelated to this analyzer.
RDW, red cell distribution width; WBC, white blood cell count.

## Discussion

### Principal findings

An uncorrected change in the NHANES hematology analyzer shifted seven blood-count measures. Two of them, RDW and WBC, rose by about 6% at that boundary and at no other, by a constant amount across their distributions. Through these two inputs, the change added 2.63 years of apparent phenotypic age acceleration, produced an apparent post-2012 surge in the biological aging of US adults, and nearly doubled the share of adults examined from 2013 onward who fell in the top phenotypic age quartile when cycles were pooled. The ratio measures followed the analytes they contain: RAR and PLR shifted, whereas SII and NLR, in which WBC cancels, did not. Two fixed corrections derived in adolescents removed 92% of the phenotypic age step and the shifts in RAR and PLR, while the same procedure applied to chemistry analytes did not.

### Comparison with other evidence

When a biochemistry platform changes, NCHS runs split-sample comparisons and publishes bridging equations [8]. The hematology transition left only a qualitative caution in the cycle documentation, because whole blood cannot be stored for later comparison [9]; a reader who followed that documentation to the letter still had no way to harmonize the data. The adolescent referent applies the reasoning NCHS itself used for hemoglobin A1c, extended to every adjacent-cycle boundary, with the form of the shift tested across percentiles and the procedure tested on chemistry analytes that did not change platform. That combination both corrects RDW and WBC and shows why the red cell indices and platelets, which shift at several boundaries, cannot be corrected with a single factor. Measurement artifacts of this size are not unique to clinical laboratory clocks: technical noise alone produces deviations of up to 9 years between replicate measurements of epigenetic clocks [14], and robust measurement is a stated prerequisite for translating any biomarker of aging [15]. The rapid growth of formulaic NHANES analyses [1] makes the gap between the documentation and the analyses more consequential.

### Implications

Because the shift is additive, rankings within a single cycle are preserved. Several published studies, however, pool cycles across 2013 and compare levels, trends or quantile groups across them. In such analyses, the post-2013 offset raises every later participant relative to every earlier one, nearly doubling the share of later participants in the top pooled quartile of phenotypic age (34.6% vs 18.9%) and of RAR (36.1% vs 17.9%), and in trend analyses it manufactures a rise in biological aging that the corrected series does not show. Studies of that kind that include measures containing RDW or WBC warrant reexamination; studies confined to one side of the boundary, or to within-cycle ranks, are not affected. A recent example illustrates both sides: an aging index derived from NHANES 2005–2018 in which RDW is the second most heavily weighted input after age, benchmarked against phenotypic age, reports discrimination that is largely insensitive to the shift, whereas its clinical thresholds, mortality nomogram and cluster-derived phenotypes rest on absolute values pooled across it; the analyzer change is not mentioned [16]. We recommend three rules for NHANES analyses spanning 2013: subtract 0.72 percentage points from RDW and 0.42 × 10³/µL from WBC for cycles from 2013–2014 onward before deriving composite measures; use ratios in which WBC cancels, such as NLR, without adjustment; and do not pool red cell indices or platelet-based measures across cycles.

### Strengths and limitations

The analysis tested every adjacent-cycle boundary rather than only the one of interest, tested the form of the correction across the distribution, and used chemistry analytes as placebo tests of the correction procedure. Several limitations should be noted. The correction rests on an adolescent referent rather than split samples, which NCHS states are not possible for this panel; a true secular change in adolescents at that boundary would bias it, although the absence of comparable changes at the other nine boundaries argues against one. Correction factors derived in adolescents are applied to adults, which assumes an additive shift that does not depend on age; the similar step across adult age groups supports this. Part of the 2011–2012 cycle was measured on the new analyzer, so the estimated changes are conservative. NHANES reports RDW and WBC to one decimal place, which limits the resolution of the percentile test. The earlier chemistry platform used in 1999–2000 is not addressed here.

### Conclusions

An uncorrected hematology analyzer change distorts blood-count measures of aging and inflammation pooled across NHANES cycles, and two fixed values correct the inputs that can be corrected. The hematology analyzer is only one of many method transitions across NHANES. As multicycle analyses multiply, researchers, package developers, reviewers and editors should verify each cycle’s documentation for every variable before pooling data.

## Data Availability

No new data were generated by this study. All data used are publicly available from the National Health and Nutrition Examination Survey (NHANES), 1999–2000 through August 2021–August 2023, conducted by the National Center for Health Statistics (https://wwwn.cdc.gov/nchs/nhanes/). The R scripts that download the data, build the analytic frame and perform all analyses, the script that draws Fig 1, and the results files underlying every number reported are available at https://github.com/TVBarreira/NHANES-hematology-analyzer-change under the MIT license.

https://wwwn.cdc.gov/nchs/nhanes/

## Supporting information

**S1 STROBE Checklist.** STROBE checklist for cross-sectional studies, with page numbers and text excerpts.

**S1 Table.** Change in each blood-count and chemistry analyte between adjacent NHANES cycles in adolescents aged 12–19 years, all 10 boundaries.

**S2 Table.** Change in the NCHS-identified blood-count measures at the analyzer boundary, by percentile of the adolescent distribution.

## Data availability

All NHANES data are publicly available from the National Center for Health Statistics (https://wwwn.cdc.gov/nchs/nhanes/). The R scripts that build the analytic frame and perform all analyses, the script that draws Fig 1, and the results files that underlie every number reported are available at https://github.com/TVBarreira/NHANES-hematology-analyzer-change under the MIT license.

## Author contributions

Conceptualization: TVB, MKA, KSH. Data curation: TVB. Formal analysis: TVB. Methodology: TVB, MKA, KSH. Software: TVB. Visualization: TVB. Writing – original draft: TVB, ECC, KKH. Writing – review & editing: TVB, KKH, ECC, MKA, KSH. TVB had full access to all the data and takes responsibility for the integrity of the data and the accuracy of the analysis.

